# Oral HPV and Dental Profiles in Mothers and Youth with or without HIV

**DOI:** 10.64898/2026.01.05.26343463

**Authors:** Anil Kumar, Oluwaseun Peter, Esosa Osagie, Jianhong Chen, Paul Akhigbe, Jia Liu, Nosakhare Lawrence Idemudia, Peter Kavecky, Ozoemene Obuekwe, Fidelis Ewenitie Eki Udoko, Nicholas F Schlecht, Yana Bromberg, Nosayaba Osazuwa-Peters, Modupe O Coker

## Abstract

**Background:** People living with HIV (PLWH) are more susceptible to persistent human papilloma virus (HPV) infection; however, data regarding oral HPV burden among youth with or without perinatal HIV exposure or infection in sub-Saharan Africa remain scarce. This study characterized how dental, immune and behavioral factors contribute to oral HPV susceptibility among youth and mothers across varying HIV exposure groups.

**Methods:** This baseline analysis leveraged data from a prospective cohort in Nigeria. Participants were categorized at recruitment as HIV-infected (HI), HIV-uninfected (HU), HIV-exposed uninfected (HEU), or HIV-unexposed uninfected (HUU). Standardized questionnaires captured behavioral data, and comprehensive dental examinations assessed DMFT (Decayed, Missing, Filled Teeth), OHIS (Oral Hygiene Index Simplified), and GIS (Gingival Inflammation Score). Oral rinse specimens were tested for oral HPV DNA (Seegene assay). Blood samples were collected from all participants for immune parameters (CD4/CD8). Multivariable regression and machine-learning approaches were used to identify key predictors across immunologic, behavioral, and oral-health domains.

**Results:** Although overall oral HPV prevalence was low, detection significantly differed across study groups. Oral HPV DNA was exclusively detected in mothers living with HIV (N=8/404) and 7 youth (N=7/671; HI = 4, HEU = 2, HUU = 1). Among youth, HPV correlated with lower CD4/CD8 ratios and poorer oral health In mothers, HPV positivity was linked to earlier sexual debut and lower CD4 counts. Machine learning models revealed distinct age-specific patterns; dental metrics and immune measures were the primary predictors in youth, outranking traditional behavioral factors whereas immune features and dental indices dominated in mothers.

**Conclusions:** Despite low prevalence, oral HPV clustered among PLWH and was strongly associated with modifiable dental indices. These findings identify oral health as a potential determinant of HPV susceptibility and underscore the importance of integrating oral health promotion within HIV care to elucidate and mitigate pathways linking oral health, immunity, and viral persistence.

## Introduction

Despite major advances in antiretroviral therapy (ART), Human Immunodeficiency Virus (HIV) infection remains a major global challenge. Of the estimated 40 million people living with HIV (PLWH) worldwide, only about three-quarters are receiving treatment, according to the *Global Burden of Diseases, Injuries and Risk Factors Study 2021*(Bassey and Miteu 2023; GBD 2021 HIV Collaborators 2024). Although the incidence of new infections has decreased over the past 20 years, overall prevalence is expected to rise through 2050 due to improved survival and population growth (Bassey and Miteu 2023; GBD 2021 HIV Collaborators 2024). The ongoing burden of HIV continues to strain health systems, impede socioeconomic development, and sustain regional disparities,(Emmanuel et al. 2024; Buchbinder and Liu 2025) particularly in sub-Saharan Africa (SSA), where Nigeria accounts for the highest number of new infections in West and Central Africa (Denny et al. 2012; Lekoane et al. 2020). These patterns reflect persistent challenges related to access to care and stigma, which underscore the need for a sustainable and equitable global plan. HIV infection also amplifies susceptibility to other sexually transmitted infections (STIs), notably human papilloma virus (HPV) (Riddell et al. 2022). HIV-induced immune dysregulation facilitates HPV acquisition and persistence, contributing to higher rates of HPV-associated malignancies despite ART-mediated viral suppression (Grulich et al. 2007).

The classification of HPV includes low-risk subtypes (causing warts) and high-risk subtypes (associated with cervical, oropharyngeal, and other anogenital cancers). Among viral causes of cancer, HPV accounts for approximately 99% of cervical cancers, 90% of anal cancers, and 70% of oropharyngeal cancers (Beachler et al. 2012; Badial et al. 2018; Bayram et al. 2025). Transmission can occur through sexual contact, skin-to-skin contact, and, although relatively rare, vertically from mother to child during childbirth (Wolf et al. 2024). Oropharyngeal cancers (OPC) can be caused by high-risk HPV, and the oral cavity serves as both a reservoir and an interface where viral persistence is influenced by oral hygiene, mucosal integrity, and salivary immune function (Kahn et al. 2015). Classic dental indices and scores could reflect inflammatory and epithelial vulnerability that may facilitate HPV colonization (Mazul et al. 2017). While oral HPV shares biological similarities with anogenital infection it tends to clear more rapidly (Beachler et al. 2012).

Despite the high burden of HIV in sub-Saharan Africa, data on oral HPV infection in the region remain exceedingly sparse, particularly among PLWH and their perinatally exposed or infected youth who may not yet be sexually active. This gap is critical because oral HPV is shaped not only by sexual behavior but also by immune function and oral health, yet these pathways remain largely unexamined in African populations. Although the mechanisms connecting oral health and oral HPV are not fully understood, poor oral hygiene and chronic oral inflammation may create conditions that promote HPV persistence and replication in the oral cavity (Giuliano et al. 2023). Existing studies link low CD4 counts and prolonged HIV duration to HPV progression, but these relationships have not been explored in the context of oral health or in youth, who represent a biologically distinct group (Beachler and D’Souza 2013; Gatechompol et al. 2020; Coker et al. 2024). As increasing numbers of children with perinatally acquired HIV survive into adolescence and adulthood, understanding their vulnerability to co-infections such as HPV is urgently needed (Croucher et al. 2013). These youth experience prolonged early-life immune dysregulation, often combined with limited access to HPV vaccination and high parental HPV prevalence, placing them at potentially heightened risk of acquiring and maintaining high-risk HPV infections (Phanuphak et al. 2019).

To address these gaps, the present study aims to characterize the burden of oral HPV among Nigerian youth and their mothers and to examine how HIV status, immune function, and oral health indicators and indices jointly influence HPV infection. By integrating immunology and dental determinants, this work aims to generate evidence essential for targeted HPV prevention and early oral cancer risk reduction within HIV care frameworks in sub-Saharan Africa.

## Methods

### Study Design and Participants

This analysis used baseline data from the *Human Papillomavirus, Human Immunodeficiency Virus, and Oral Microbiota Interplay in Nigerian Youth (HOMINY)* study, a prospective cohort at the University of Benin Teaching Hospital, Nigeria (Osagie et al. 2025). Between October 2022 and May 2023, youth aged 9–18 years and their mothers were enrolled. Youth were classified as HIV-infected (HI), HIV-exposed uninfected (HEU), or HIV-unexposed uninfected (HUU), and mothers as HIV-infected (HI) or uninfected (HU). Youth in the HI and HEU groups were age- and sex-matched to participants in the HUU group. Participant recruitment was conducted by trained healthcare professionals who approached eligible youth–mother pairs during routine clinic visits. Research staff provided standardized written and verbal explanations of study procedures, risks, and benefits. Mothers provided informed consent for themselves and their children. Individuals who indicated interest and met eligibility criteria were enrolled into the cohort.

To account for an expected 15% loss to follow-up, we aimed to enroll 230 youth–mother pairs in each group (HI, HEU, and HUU), for a total target of 690 participants. The HUU group served as the reference for comparisons. With an effective sample of about 600 participants after attrition, the study maintained approximately 80% power to detect incidence rate ratios around 1.2 in Poisson models (α=0.05, two-sided). Power estimates were similar when adjusting for additional covariates.

### Clinical and Behavioral Data Collection

Standardized questionnaires collected sociodemographic information (age, sex, education, employment, marital status, and number of pregnancies and number of children) and behavioral variables (tobacco/alcohol use, current consumption). Sexual health variables included age at sexual debut, number of lifetime sexual partners, sexual practices such as use of condom, and HPV awareness and vaccination history. Comprehensive oral examinations were performed by calibrated dental clinicians following WHO protocols. Caries experience was measured using the Decayed, Missing, and Filled Teeth (DMFT) index (range, 0–28), while the Pulpal Involvement, Ulceration, Fistula, and Abscess (PUFA) index (range, 0–28) captured sequelae of untreated decay. Oral hygiene was evaluated using the Oral Hygiene Index–Simplified (OHIS; range, 0–6) and categorized as good, fair, or poor. Gingival status was recorded via the Gingival Inflammation Score (GIS; range, 0–3), reflecting local inflammatory response. All indices were assessed under standardized illumination and sterile conditions, providing quantitative markers of oral disease burden relevant to HPV susceptibility. Biological samples collected included whole blood (for CD4 and CD8 lymphocyte counts), and oral rinse specimens (for HPV DNA detection and genotyping).

### HPV Sample Collection and Testing

HPV testing was conducted at two laboratories, Inqaba Biotechnical Industries (Nigeria) and Roswell Park Comprehensive Cancer Center (NY, USA) using identical protocols. DNA from oral rinse samples was extracted with the Zymo Quick-DNA Miniprep Kit and tested with the Seegene Allplex HPV28 assay, detecting 19 high-risk and 9 low-risk genotypes (Bell et al. 2023). A sample was deemed HPV-positive if amplification was reproducible with a Ct ≤ 35 or reproducible at higher Ct values.

### Statistical Analysis

Descriptive statistics summarized participant characteristics by HIV and HPV status. Continuous variables were compared using t-tests or ANOVA (Kruskal–Wallis when non-normal); categorical variables used χ ² or Fisher’s exact tests. To identify variables most strongly associated with oral HPV detection, we applied Random Forest (RF) models separately for mothers and youth, using all available sociodemographic, behavioral, immunologic, and oral-health variables.^23^ Briefly, an RF model was first built using each predictor to remove its association with the outcome. We tested robustness using the Boruta model for predicting the association of predictors to oral HPV.^24^ Additional details describing these models can be found in supplementary materials.

### Ethical Approval

The institutional review boards at Rutgers State University (Pro2022000949) and the UBTH, Benin City (ADM/E22/A/VOL. VII/14813674), Nigeria, approved this study and will be informed of the study findings in a timely manner.

### Reporting Guidelines

This study adhered to the STROBE guidelines for reporting observational cohort studies.

## Results

Baseline demographic, socioeconomic, and immunologic profiles of youths and mothers are summarized in **Table 1 and 2**. The youth cohort included 671 participants, evenly distributed across HI (N=226), HEU (N=219), and HUU (N=226) groups. Most youths were delivered vaginally, though the HEU group were more likely to have been delivered via cesarean section compared to other groups (p=0.002). Educational attainment reflected appropriate age levels with most of the youth participants having completed either primary or junior secondary school. Health behavior data indicated very low engagement in risky behaviors. Almost no youth reported tobacco use or regular alcohol consumption.

**Table 1.**
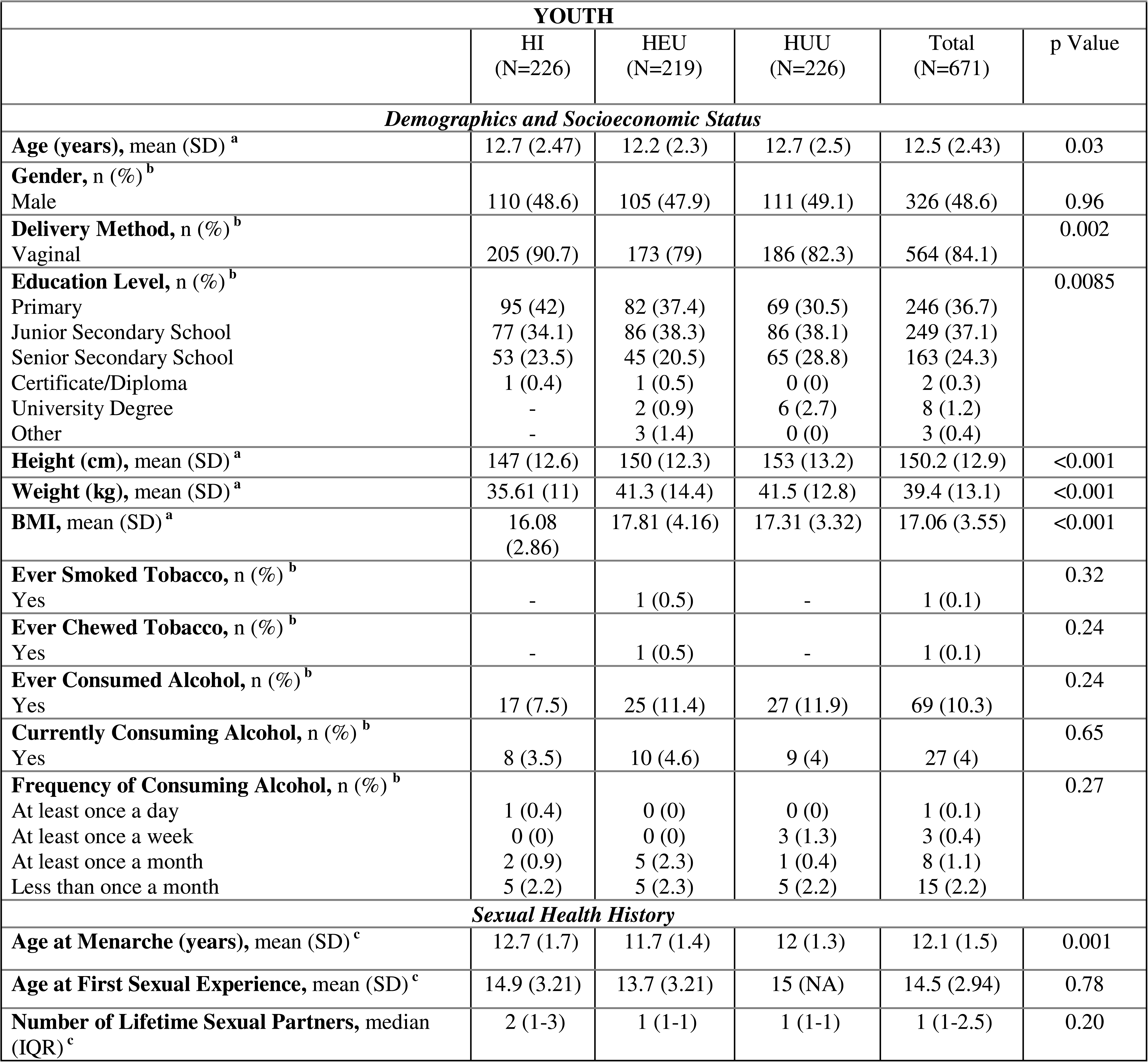
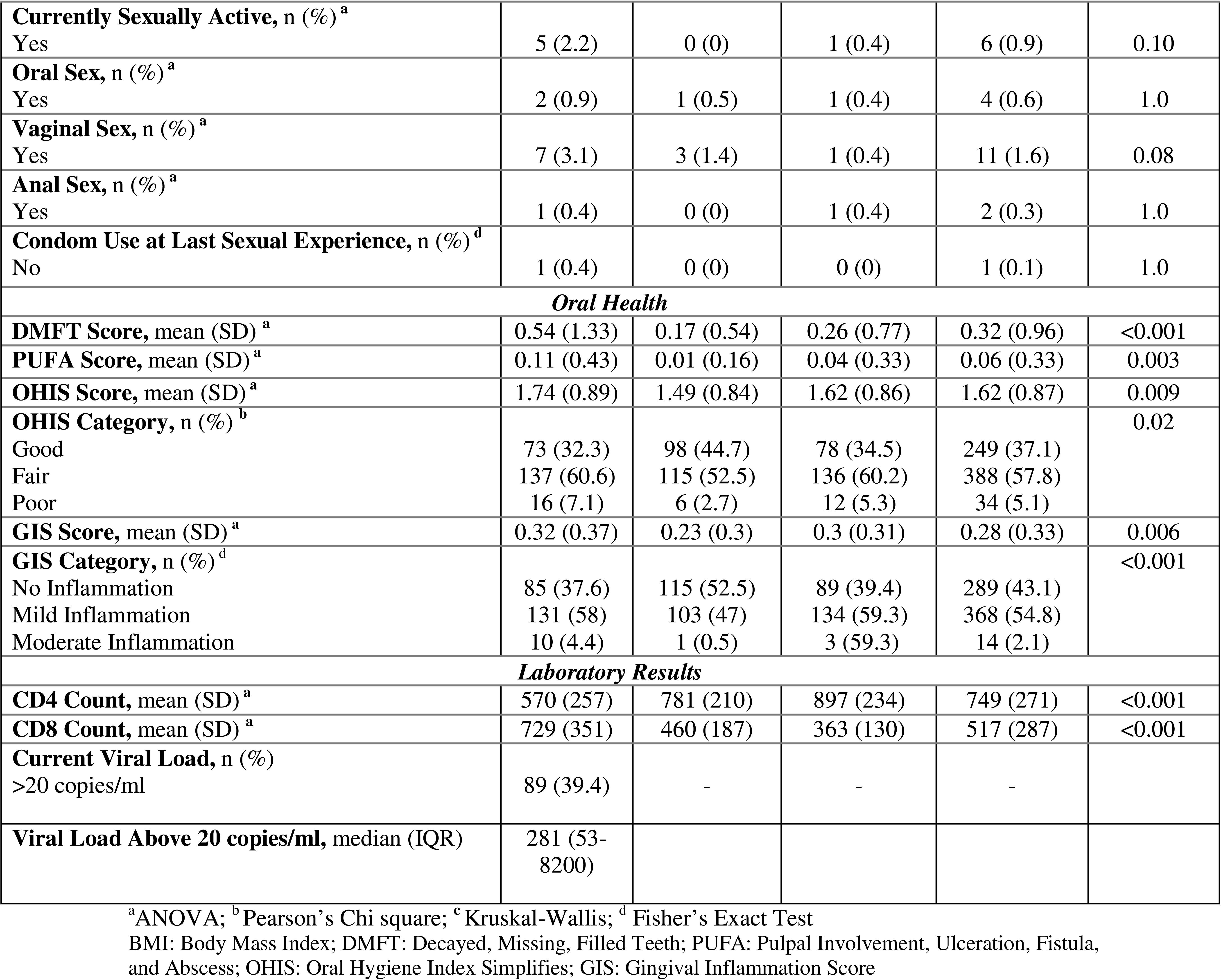
Baseline demographic, behavioral, immunologic, and oral health characteristics of youth participants, stratified by HIV status.

| **YOUTH** |  |  |  |  |  |
| --- | --- | --- | --- | --- | --- |
|  | HI<br>(N=226) | HEU<br>(N=219) | HUU<br>(N=226) | Total<br>(N=671) | p Value |
| *Demographics and Socioeconomic Status* |  |  |  |  |  |
| **Age (years), mean (SD) <sup>a</sup>** | 12.7 (2.47) | 12.2 (2.3) | 12.7 (2.5) | 12.5 (2.43) | 0.03 |
| **Gender, n (%) <sup>b</sup>** |  |  |  |  |  |
| Male | 110 (48.6) | 105 (47.9) | 111 (49.1) | 326 (48.6) | 0.96 |
| **Delivery Method, n (%) <sup>b</sup>** |  |  |  |  | 0.002 |
| Vaginal | 205 (90.7) | 173 (79) | 186 (82.3) | 564 (84.1) |  |
| **Education Level, n (%) <sup>b</sup>** |  |  |  |  | 0.0085 |
| Primary | 95 (42) | 82 (37.4) | 69 (30.5) | 246 (36.7) |  |
| Junior Secondary School | 77 (34.1) | 86 (38.3) | 86 (38.1) | 249 (37.1) |  |
| Senior Secondary School | 53 (23.5) | 45 (20.5) | 65 (28.8) | 163 (24.3) |  |
| Certificate/Diploma | 1 (0.4) | 1 (0.5) | 0 (0) | 2 (0.3) |  |
| University Degree | - | 2 (0.9) | 6 (2.7) | 8 (1.2) |  |
| Other | - | 3 (1.4) | 0 (0) | 3 (0.4) |  |
| **Height (cm), mean (SD) <sup>a</sup>** | 147 (12.6) | 150 (12.3) | 153 (13.2) | 150.2 (12.9) | <0.001 |
| **Weight (kg), mean (SD) <sup>a</sup>** | 35.61 (11) | 41.3 (14.4) | 41.5 (12.8) | 39.4 (13.1) | <0.001 |
| **BMI, mean (SD) <sup>a</sup>** | 16.08<br>(2.86) | 17.81 (4.16) | 17.31 (3.32) | 17.06 (3.55) | <0.001 |
| **Ever Smoked Tobacco, n (%) <sup>b</sup>** |  |  |  |  | 0.32 |
| Yes | - | 1 (0.5) | - | 1 (0.1) |  |
| **Ever Chewed Tobacco, n (%) <sup>b</sup>** |  |  |  |  | 0.24 |
| Yes | - | 1 (0.5) | - | 1 (0.1) |  |
| **Ever Consumed Alcohol, n (%) <sup>b</sup>** |  |  |  |  | 0.24 |
| Yes | 17 (7.5) | 25 (11.4) | 27 (11.9) | 69 (10.3) |  |
| **Currently Consuming Alcohol, n (%) <sup>b</sup>** |  |  |  |  | 0.65 |
| Yes | 8 (3.5) | 10 (4.6) | 9 (4) | 27 (4) |  |
| **Frequency of Consuming Alcohol, n (%) <sup>b</sup>** |  |  |  |  | 0.27 |
| At least once a day | 1 (0.4) | 0 (0) | 0 (0) | 1 (0.1) |  |
| At least once a week | 0 (0) | 0 (0) | 3 (1.3) | 3 (0.4) |  |
| At least once a month | 2 (0.9) | 5 (2.3) | 1 (0.4) | 8 (1.1) |  |
| Less than once a month | 5 (2.2) | 5 (2.3) | 5 (2.2) | 15 (2.2) |  |
| *Sexual Health History* |  |  |  |  |  |
| **Age at Menarche (years), mean (SD) <sup>c</sup>** | 12.7 (1.7) | 11.7 (1.4) | 12 (1.3) | 12.1 (1.5) | 0.001 |
| **Age at First Sexual Experience, mean (SD) <sup>c</sup>** | 14.9 (3.21) | 13.7 (3.21) | 15 (NA) | 14.5 (2.94) | 0.78 |
| **Number of Lifetime Sexual Partners, median (IQR) <sup>c</sup>** | 2 (1-3) | 1 (1-1) | 1 (1-1) | 1 (1-2.5) | 0.20 |
| **Currently Sexually Active, n (%)**<sup>a</sup> |  |  |  |  |  |
| Yes | 5 (2.2) | 0 (0) | 1 (0.4) | 6 (0.9) | 0.10 |
| **Oral Sex, n (%)**<sup>a</sup> |  |  |  |  |  |
| Yes | 2 (0.9) | 1 (0.5) | 1 (0.4) | 4 (0.6) | 1.0 |
| **Vaginal Sex, n (%)**<sup>a</sup> |  |  |  |  |  |
| Yes | 7 (3.1) | 3 (1.4) | 1 (0.4) | 11 (1.6) | 0.08 |
| **Anal Sex, n (%)**<sup>a</sup> |  |  |  |  |  |
| Yes | 1 (0.4) | 0 (0) | 1 (0.4) | 2 (0.3) | 1.0 |
| **Condom Use at Last Sexual Experience, n (%)**<sup>d</sup> |  |  |  |  |  |
| No | 1 (0.4) | 0 (0) | 0 (0) | 1 (0.1) | 1.0 |
| **Oral Health** |  |  |  |  |  |
| **DMFT Score, mean (SD)**<sup>a</sup> | 0.54 (1.33) | 0.17 (0.54) | 0.26 (0.77) | 0.32 (0.96) | <0.001 |
| **PUFA Score, mean (SD)**<sup>a</sup> | 0.11 (0.43) | 0.01 (0.16) | 0.04 (0.33) | 0.06 (0.33) | 0.003 |
| **OHIS Score, mean (SD)**<sup>a</sup> | 1.74 (0.89) | 1.49 (0.84) | 1.62 (0.86) | 1.62 (0.87) | 0.009 |
| **OHIS Category, n (%)**<sup>b</sup> |  |  |  |  | 0.02 |
| Good | 73 (32.3) | 98 (44.7) | 78 (34.5) | 249 (37.1) |  |
| Fair | 137 (60.6) | 115 (52.5) | 136 (60.2) | 388 (57.8) |  |
| Poor | 16 (7.1) | 6 (2.7) | 12 (5.3) | 34 (5.1) |  |
| **GIS Score, mean (SD)**<sup>a</sup> | 0.32 (0.37) | 0.23 (0.3) | 0.3 (0.31) | 0.28 (0.33) | 0.006 |
| **GIS Category, n (%)**<sup>d</sup> |  |  |  |  | <0.001 |
| No Inflammation | 85 (37.6) | 115 (52.5) | 89 (39.4) | 289 (43.1) |  |
| Mild Inflammation | 131 (58) | 103 (47) | 134 (59.3) | 368 (54.8) |  |
| Moderate Inflammation | 10 (4.4) | 1 (0.5) | 3 (59.3) | 14 (2.1) |  |
| **Laboratory Results** |  |  |  |  |  |
| **CD4 Count, mean (SD)**<sup>a</sup> | 570 (257) | 781 (210) | 897 (234) | 749 (271) | <0.001 |
| **CD8 Count, mean (SD)**<sup>a</sup> | 729 (351) | 460 (187) | 363 (130) | 517 (287) | <0.001 |
| **Current Viral Load, n (%)** |  |  |  |  |  |
| >20 copies/ml | 89 (39.4) | - | - | - |  |
| **Viral Load Above 20 copies/ml, median (IQR)** | 281 (53-8200) |  |  |  |  |

<sup>a</sup>ANOVA; <sup>b</sup> Pearson's Chi square; <sup>c</sup> Kruskal-Wallis; <sup>d</sup> Fisher's Exact Test
BMI: Body Mass Index; DMFT: Decayed, Missing, Filled Teeth; PUFA: Pulpal Involvement, Ulceration, Fistula, and Abscess; OHIS: Oral Hygiene Index Simplifies; GIS: Gingival Inflammation Score

Of the 404 mothers enrolled (275 HI and 129 HU), mothers living with HIV (MLWH) were slightly older, had lower educational attainment, higher unemployment, and lower mean CD4 counts than HU mothers (p<0.05; **Table 2**). Health behavior indicators showed that smoking and tobacco chewing were extremely rare in this population, with nearly all mothers reporting no history of either behavior (2 of 404). Alcohol consumption, however, was relatively more common. The mean age at first sexual experience was around 19.6 years (p<0.001), and more than 84% reported being sexually active at the time of the study. Sexual debut occurred earlier among mothers in the HI group when compared to mothers in the HU group, suggesting a greater window of potential exposure to sexually transmitted infections (p <0.001). Condom use at last sexual encounter was low (63.6%) and all mothers reported vaginal sex as their primary sexual practice, while oral and anal sex were reported infrequently (12.1% and 5.2% respectively). Awareness of HPV and the HPV vaccine was notably limited (**Table 2**).

**Table 2.**
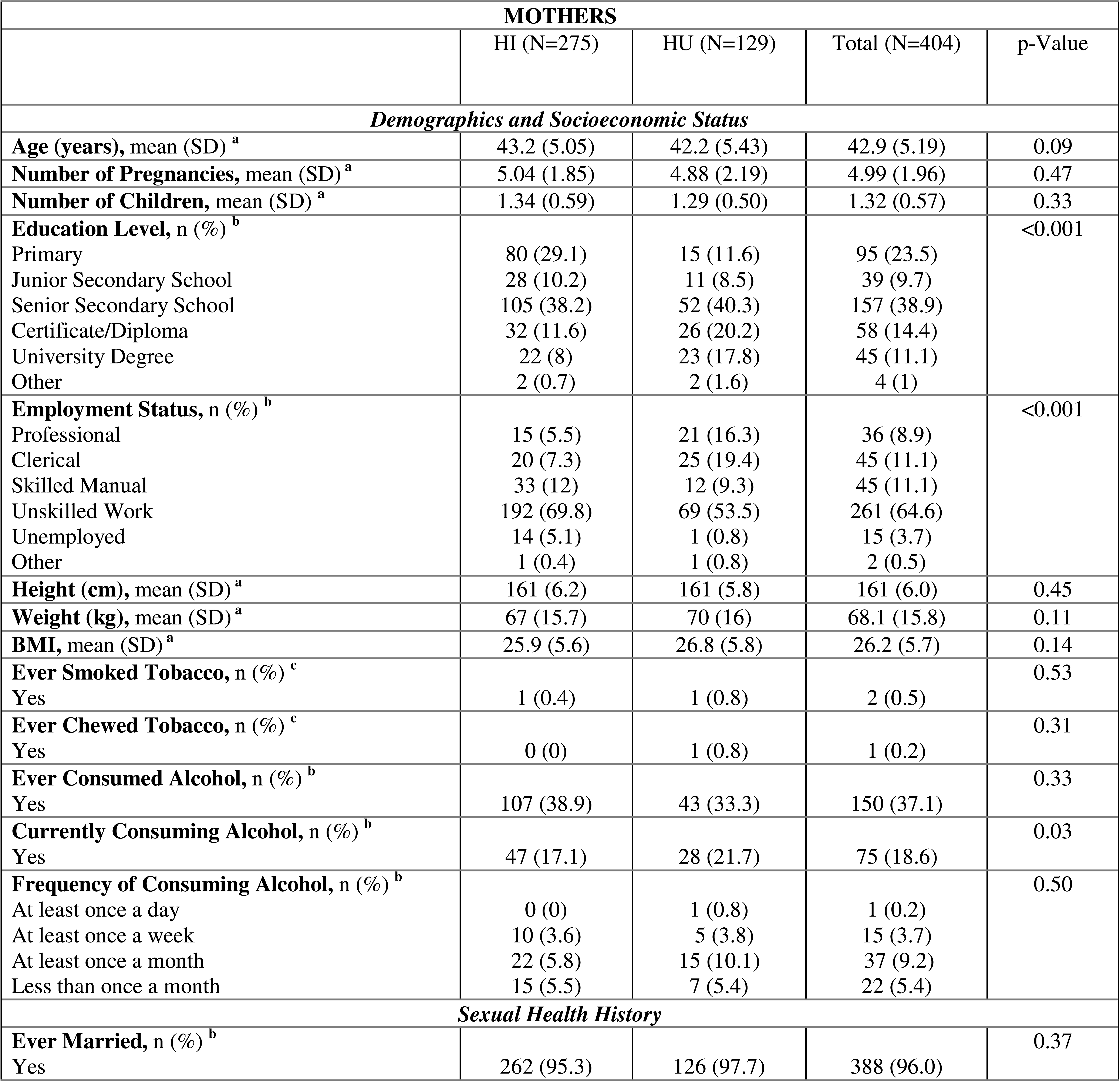
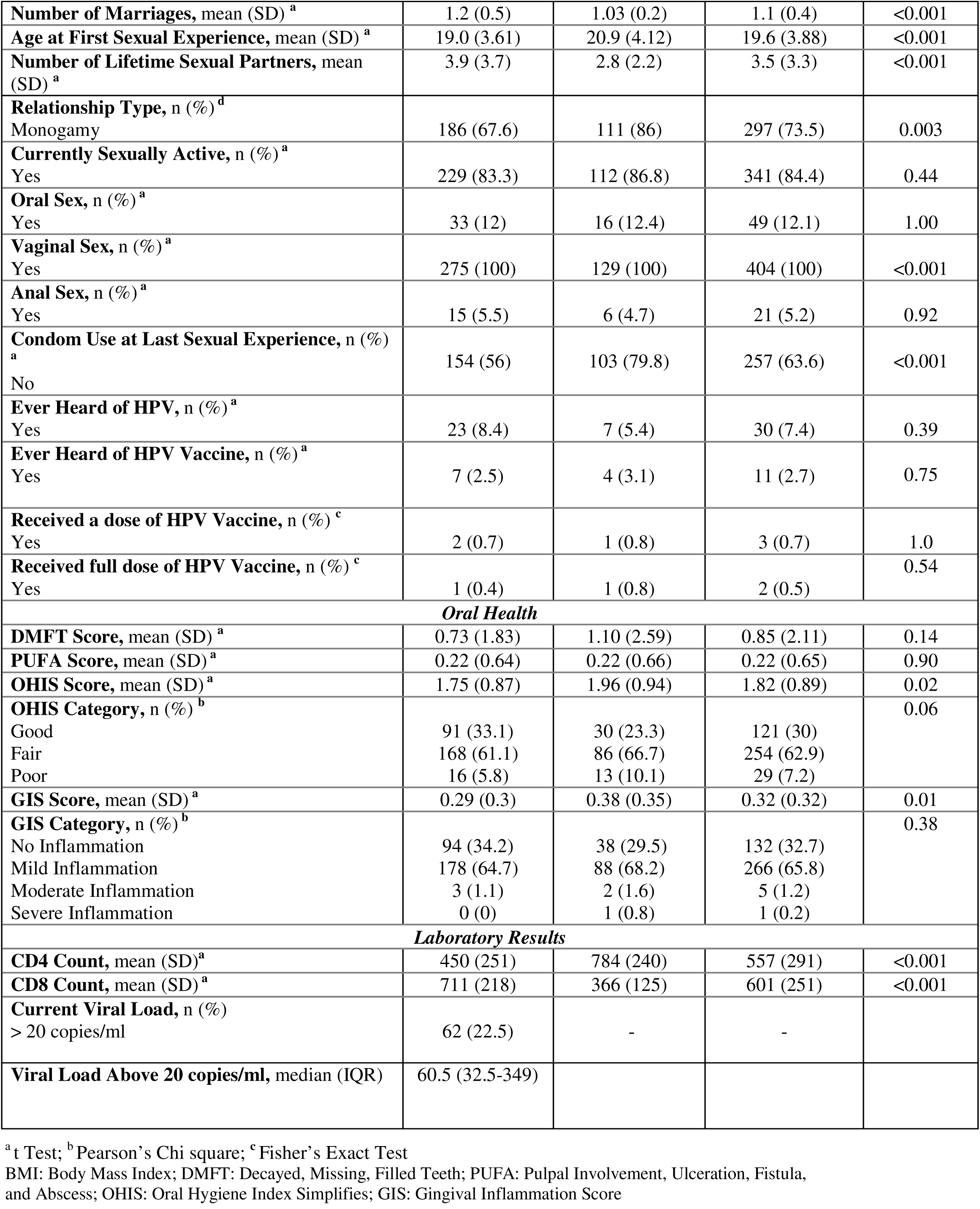
Baseline demographic, behavioral, immunologic, and oral health characteristics of mother participants, stratified by HIV status.

Oral HPV was detected across all youth groups (HI, HEU, and HUU) but exclusively among MLWH (**Figure 1**). In this cohort, we identified the presence of 12 distinct human papillomavirus (HPV) DNA strains, including HPV types 11, 16, 33, 35, 39, 40, 56, 58, 59, 60, 61, and 70. HPV16 DNA was the most prevalent strain detected in 20% of participants. Most participants (73%) tested positive for high-risk HPV DNA strains.

**Figure 1.**
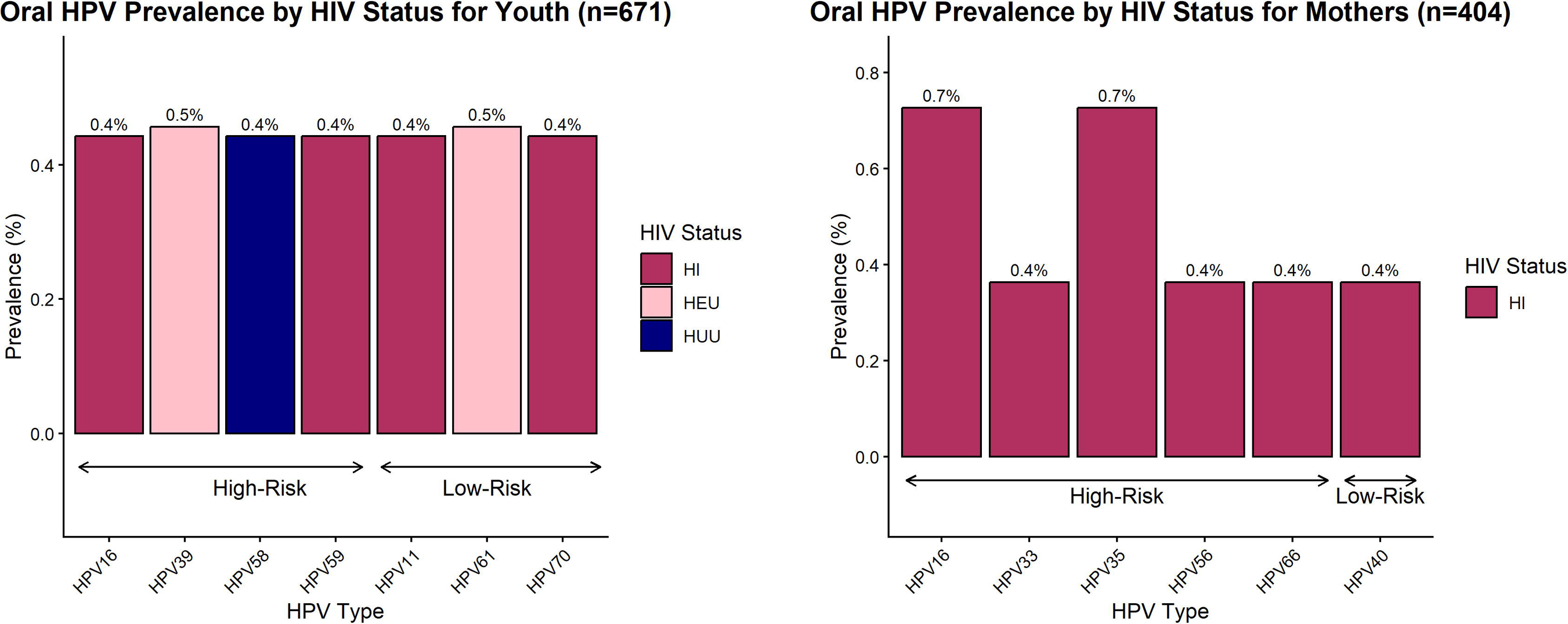
Prevalence of oral HPV DNA stratified by HIV status among (A) youth and (B) mother participants.

**Figure 2.**
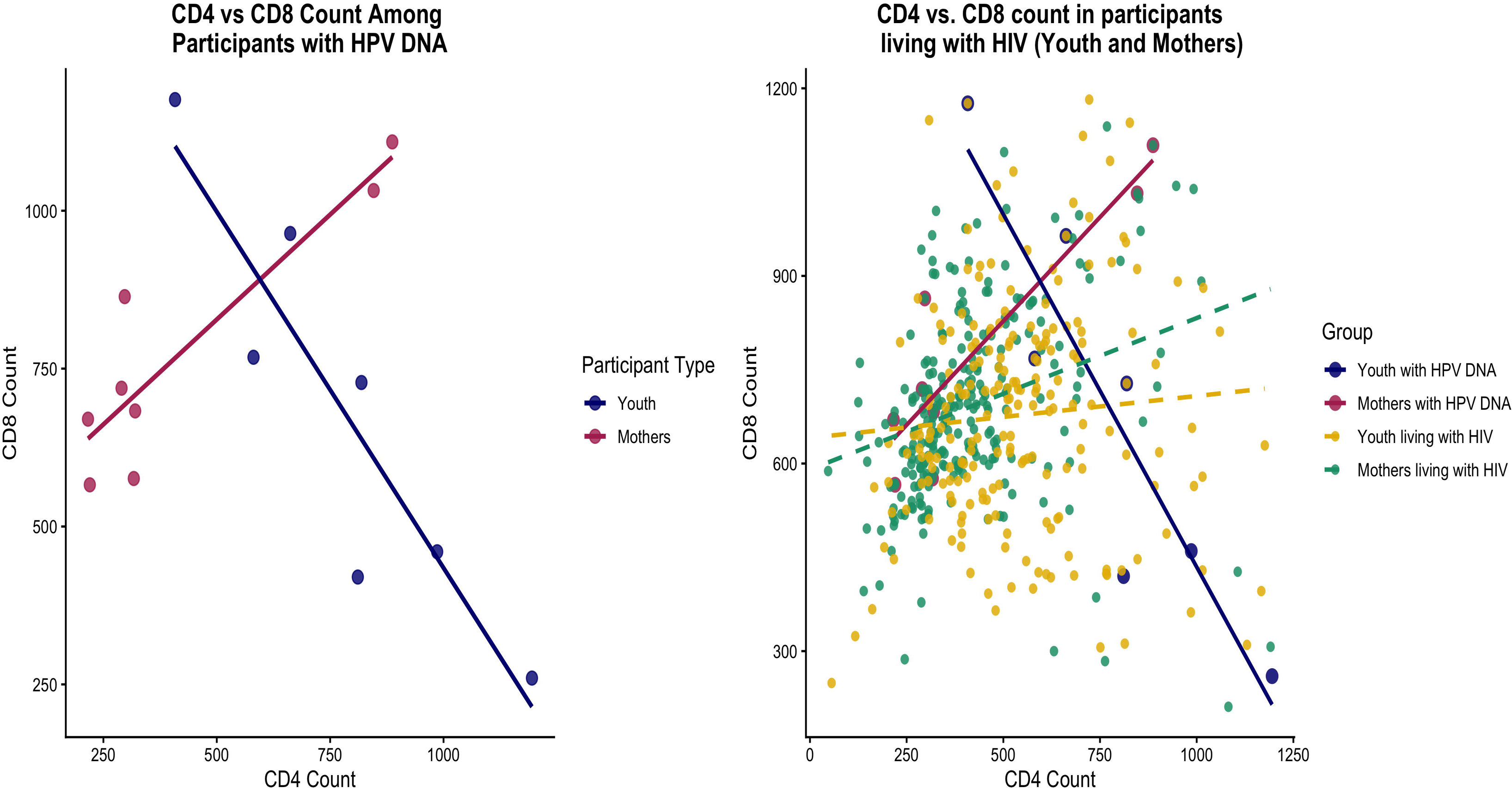
CD4 and CD8 T-cell counts by HPV and HIV status. Each point represents an individual participant. **2A.** Among HPV-positive participants, youth exhibit an inverse CD4–CD8 relationship, while mothers show a positive association. **2B.** Among participants living with HIV, regression lines depict CD4–CD8 relationships stratified by HPV DNA status (solid = HPV-positive; dashed = HPV-negative). Participants with CD8 counts >1200 cells/mm³ were excluded for visualization clarity.

HPV DNA was detected in 7/671 (1.0%) youths and 8/404 (2.0%) mothers. HPV DNA was detected mainly in youth perinatally HIV-exposed with or without perinatal acquisition (86%), comprising four HI, two HEU, and one HUU participant (**Figure 1A**). Among mothers, HPV DNA was detected exclusively in MLWH. Notably, none of the youth with HPV DNA reported sexual activity, which is consistent with the high prevalence observed among mothers in the cohort, as seven of eight mothers carried high-risk HPV DNA. Furthermore, the strong association between high-risk HPV infection and HIV status was notable, with nine of the eleven high-risk infections occurring in PLWH.

Among youths, 86% of HPV-positive cases were perinatally HIV-exposed or infected (four HI, two HEU, one HUU), and notably, none reported sexual activity. All mothers with detectable HPV DNA belonged to the HI group and the majority (7 out of 8) of them carried high-risk types. Overall, nine of eleven high-risk infections occurred in PLWH.

Immune profiling demonstrated distinct patterns associated with HPV detection in both youth and mothers. Among youth living with HIV, those with detectable HPV DNA showed slightly higher CD4 counts (780 vs 570 cells/mm³) and lower CD8 counts (682 vs 729 cells/mm³) compared to their counterparts. In contrast, MLWH and detectable HPV DNA exhibited lower CD4 counts (424 vs 451 cells/mm³) and higher CD8 counts (777 vs 711 cells/mm³), consistent with immune activation typical of chronic HIV infection **(Supplementary Figure 4)**. Specifically, all seven youths had a DMFT score of 0 and no PUFA involvement, while most showed “good” OHIS (85.7%) and minimal gingival inflammation. In contrast, mothers with oral HPV showed poorer oral hygiene, with only one having a DMFT score above 2 and the majority falling into the “fair” OHIS category **(Supplementary Figure 3)**. Their PUFA scores, although low, were still higher on average than those of youth, and 75% had signs of mild gingival inflammation. This pattern suggests that despite relatively healthy dentition, oral HPV can occur in the absence of overt disease, potentially mediated by immune dysregulation.

In contrast, mothers with detectable HPV DNA demonstrated a higher oral disease burden, including fair-to-poor OHIS in most cases, elevated PUFA scores, and mild gingival inflammation in 75% of participants. These findings highlight the cumulative influence of oral microbial ecology, mucosal inflammation, and systemic immune compromise in promoting viral persistence among PLWH.

Sexual histories differed by HIV infection. MLWH reported a higher lifetime number of sexual partners per year than HU mothers (3.9 and 2.8 respectively, p <0.001) **(Supplementary Figure 2)**. However, when stratified by HPV status, no difference was observed in partner rates between mothers who had HPV DNA vs. without HPV DNA. These findings indicate that while HIV infection is associated with higher partner acquisition rates, HPV detection in this cohort was not explained by variation in reported partner number.

We also conducted variable importance analyses via random forests to identify contrasting patterns for youths and mothers (**Figure 3**). Behavioral and contextual factors emerged as the most important predictors of HPV status in mothers. Variables related to sexual history including the number of lifetime sexual partners, age at sexual debut, average number of sexual partners per year, CD8 count, GIS category, and demographic variables like employment status, parity, and age were ranked higher as variables of importance in predicting oral HPV. In youths, immunologic and oral health variables, CD8 counts, OHIS, DMFT, age, and education status were the strongest predictors of oral HPV (**Figure 3A**). Behavioral factors like alcohol and tobacco consumption, DMFT, and sexual behavior (condom use, sexual partners) had little predictive value, consistent with the very low prevalence of sexual activity in this age group.

**Figure 3.**
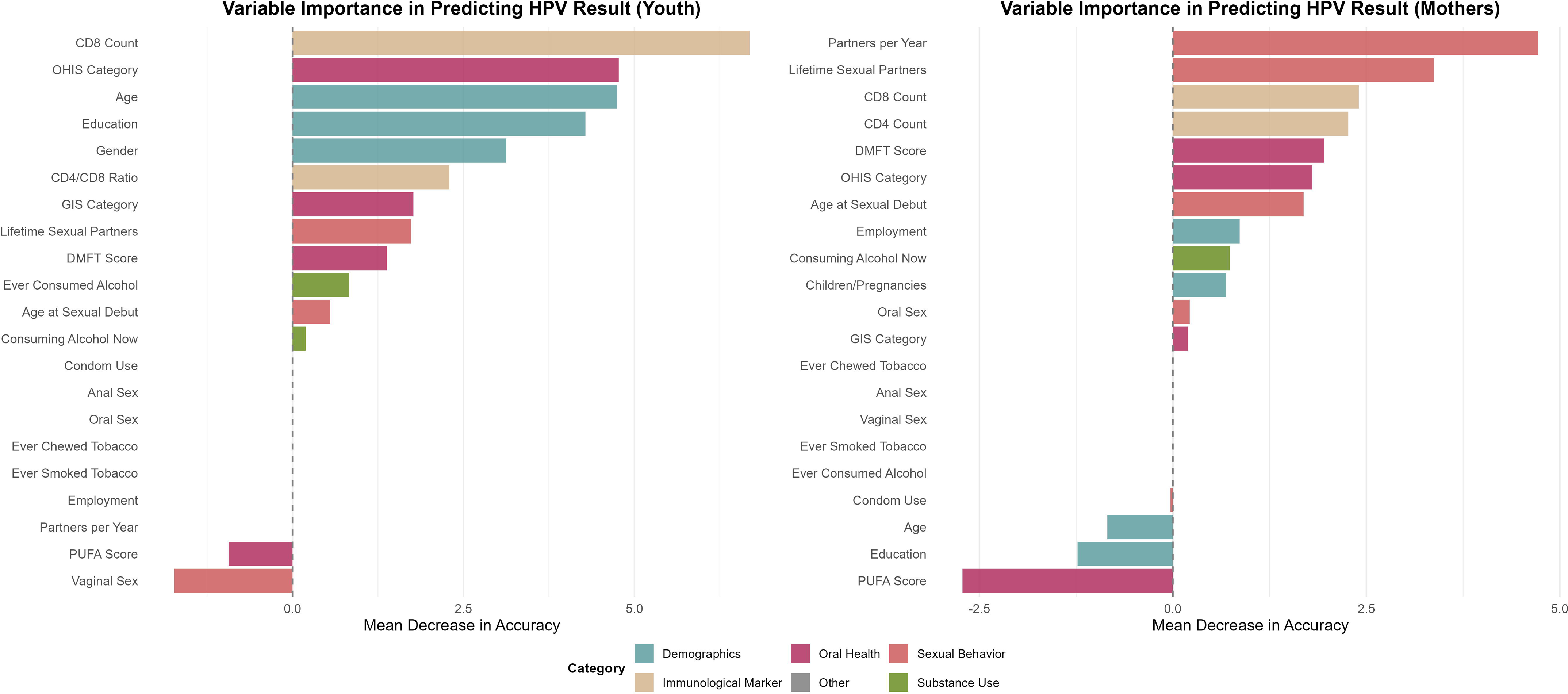
Variable importance from random forest models predicting oral HPV DNA detection. Bars represent predictors ranked by their contribution to model accuracy (mean decrease in accuracy). **3A.** In youth, top predictors included immunologic markers (CD8 count, CD4:CD8 ratio) and oral-health indices (OHIS). **3B.** In mothers, sexual-behavior factors (lifetime partners, partners per year, age at sexual debut) were strongest predictors, followed by immune and oral-health variables. Bar colors denote variable domains (demographic, oral health, sexual behavior, immunologic, substance use, other).

## Discussion

To our knowledge, this is the first study to characterize oral HPV DNA from oral rinses collected from a large, well-defined cohort of youth and their mothers in Nigeria, including both perinatally HI and HEU populations. Although the overall prevalence of oral HPV was low (1% among youth and 2% among mothers), these estimates are consistent with reports from non-sexually active or pediatric cohorts, where prevalence typically ranges from 1–5% (Castellsagué et al. 2009; Gillison et al. 2012; Chung et al. 2014; Giuliano et al. 2023). The relatively low prevalence observed in our HIV-infected youth likely reflect early and sustained ART exposure, enabling many to reach adolescence with partial immune reconstitution that reduces susceptibility to new viral acquisition (Kahn et al. 2015; Moscicki et al. 2016; Visalli et al. 2021; Coker et al. 2024)

Importantly, all HPV-positive mothers were living with HIV, and most harbored high-risk genotypes such as HPV-16 and HPV-35, which are associated with oropharyngeal and cervical cancers (Mcharo et al. 2021; Coker et al. 2024). The presence of high-risk genotypes in a subset of mothers signals the need for enhanced oral cancer risk assessment within HIV care, particularly in low-resource settings where routine dental screening is uncommon (Riddell et al. 2022). The presence of high-risk genotypes underscores the potential risk of malignant transformation, particularly in PLWH (Beachler and D’Souza 2013; Bayram et al. 2025) (**Figure 1**). Although oral HPV prevalence in youth was low, its detection in non-sexually active adolescents is notable (Petca et al. 2020; Syrjänen et al. 2021). The predominance of high-risk HPV in youth living with HIV or perinatally exposed is notable, given their smaller representation in the cohort. These findings raise the possibility of vertical, perinatal, or non-sexual horizontal transmission pathways, which have been described in prior pediatric HPV studies (Moscicki et al. 2014; Gatechompol et al. 2020). Previous studies have also reported prevalence of HPV in the oral mucosa of neonates, children and adolescents exposed to but not infected with HIV (Syrjänen et al. 2021). Similar reports have described HPV DNA in cord blood, placental tissue, and breast milk, supporting the plausibility of perinatal and postnatal transmission (Freitas et al. 2013). The observation that HPV was present in youth living with HIV or perinatally exposed, despite their smaller representation in the cohort, suggests that immunologic vulnerability, rather than behavioral exposure may be a key driver of early detection.

The dental component of these findings is especially relevant. Poor oral hygiene, dental caries, and gingival inflammation may create a pro-inflammatory environment that promotes viral entry or persistence in the oral mucosa. Although the mechanisms linking oral health to oral HPV infection are not fully understood, chronic oral inflammation may compromise epithelial integrity, alter salivary defenses, and increase susceptibility to HPV. In our cohort, youth with HPV tended to have poorer oral hygiene scores or higher caries burden, supporting the plausibility of oral health as a biological cofactor in early HPV acquisition. This aligns with increasing recognition that oral disease may contribute to the vulnerability of immunocompromised populations.

Our findings highlight that, for mothers, HPV infection may be the cumulative result of prolonged sexual exposure across multiple partners, compounded by HIV-related immune compromise (Massad et al. 2014; Megersa et al. 2023; del Pino et al. 2024). Coutlée et al. have similarly demonstrated that sexual behaviors, particularly the number of sexual partners, are strongly associated with oral HPV infection (Coutlée et al. 1997). The distinct risk pattern observed in youth and mothers suggests that, in youth, HPV detection may be driven less by behavioral exposure and more by immunologic vulnerability compared to the combined influence of immune compromise and cumulative sexual exposure observed in adult women (Sripan et al. 2024).

The role of vaccination in protecting against oral HPV in children is evolving (Mehanna et al. 2019). Nevertheless, evidence shows that maximum protection, regardless of site, is achieved when the vaccine is administered prior to viral exposure, ideally before sexual debut (Ardekani et al. 2022). In our study, mothers with HPV DNA reported earlier sexual debut than the overall maternal cohort (mean age 16 vs 19 years), suggesting longer potential exposure to sexually transmitted infections. These findings echo the need to integrate HPV vaccination into routine childhood immunization programs, integrate HPV education into HIV care, and develop tailored prevention strategies for children perinatally exposed to HIV who may face unique immunologic vulnerabilities (Ardekani et al. 2022). Lessons from previous studies point to HPV vaccination that aims to reduce infection rates in future generations and this should be framed as a preventive health measure, not as an endorsement of early or risky sexual activity (Ardekani et al. 2022). Extensive research efforts to reduce the burden of HPV-related conditions have resulted in the development licensed vaccines, along with several additional candidates currently in advanced stages of clinical evaluation (Kombe Kombe et al. 2021). The future goal should be make these vaccines available in limited resource settings as well as increasing the awareness to improve uptake.

There were several limitations. The cross-sectional design limits inferences about HPV persistence or clearance, and the low number of HPV-positive cases reduced statistical power for detecting risk factor associations. Self-reported behaviors, including sexual activity, may be underreported. Periodontal health was not clinically assessed with probing depth or radiographs; instead, proxy indices such as gingival inflammation and oral hygiene were used, which may underestimate subclinical disease. Future directions should incorporate microbiome and immunologic markers that could more precisely define pathways linking oral health to HPV susceptibility. Longitudinal follow-up of this cohort will be critical to evaluate persistence, clearance, and the role of the oral microbiota. Despite these limitations, the study’s strengths include its large cohort size, dual-site laboratory validation, and integration of behavioral, immunologic, and dental data. Machine-learning approaches provided complementary evidence by consistently identifying oral hygiene and dental disease burden as key correlations among youth.

Our findings emphasize that the oral environment plays a biologically meaningful role in HPV susceptibility. Dental indices capturing caries burden, oral hygiene, and gingival inflammation provide quantifiable markers of mucosal vulnerability and local immune function, offering a scientific basis for integrating oral health evaluation into HPV prevention frameworks, especially for PLWH. Strengthening the oral health of individuals living with or exposed to HIV is therefore not ancillary but essential. Routine dental screening, early management of caries and gingival inflammation, and targeted oral hygiene interventions could meaningfully reduce the biological conditions that favor HPV persistence. Integrating these practices into HIV care transforms the dental visit into an opportunity for early detection of viral susceptibility and prevention of downstream malignant transformation. In populations already facing immune challenges, optimizing oral health may represent one of the most practical, scalable, and impactful strategies to mitigate HPV-related disease risk.

## Funding Acknowledgement

This work was supported by the National Institute of Health/National Institute of Dental Craniofacial Research - (NIH/NIDCR R01DE032216; R01DE028154). The funder had no role in the study design, collection, analysis, interpretation of the data, and manuscript writing.

## Data Availability

All data produced in the present study are available upon reasonable request to the authors.

## Acknowledgements

The authors are grateful to the women, children/adolescents and families who made this study possible and to the dedicated study staff at the University of Benin Teaching Hospital and the Institute of Human Virology, Nigeria. Special thanks to all HOMINY Study Team members, including Lydia Esena-Obaweiki, Promise Olumefun, Toluwalope Gbolahan, Isioma Mayor-Nwakwuribe, Alex Olanipekun, Amara Godwins, Modupe Kadiri, Daniel Oakhu, Matthew Imoe-Auweokha, Jessica Kubeyinje, Daniel Anonyai, Erika Juhlin, and Dr. Uwagboe Odigie, for their invaluable contributions to participant engagement and recruitment, as well as sample and data collection and analysis. The authors also appreciate the dedicated efforts of the study dentists—Drs. Nneka Chukwumah, Stanley Iyorzor, Oseriemen Akhigbemen, Timothy Ahworegba, Gbenga Ojuola, and Sunday Adesiyan—for their time and expertise in conducting clinical examinations during study visits. Many thanks to Jianhong Chen and the team at Inqaba Biotec West Africa for their support in laboratory management, sample processing, and HPV DNA detection and subtyping. The de-identified datasets generated and analyzed during this study are not publicly available due to ethical and regulatory restrictions associated with human participant data, including protections for sensitive health information. This prohibits open public posting of the dataset. However, data may be made available upon request to the corresponding author, contingent on IRB approval and completion of appropriate data-use agreements.

## Author Contribution Statement

Anil Kumar contributed to analysis and interpretation, drafted and critically revised the manuscript. Oluwaseun Peter contributed to analysis and interpretation and critically revised the manuscript.

Esosa Osagie contributed to conception, design, analysis, interpretation, acquisition, and critically revised the manuscript.

Jianhong Chen contributed to analysis and critically revised the manuscript.

Paul Akhigbe contributed to conception, design, analysis, interpretation, and critically revised the manuscript.

Jia Liu contributed to analysis and interpretation and critically revised the manuscript.

Nosakhare Lawrence Idemudia contributed to conception, design, analysis, interpretation, and critically revised the manuscript.

Peter Kavecky Chen contributed to interpretation and critically revised the manuscript.

Ozoemene Obuekwe contributed to conception, design, analysis, interpretation, and critically revised the manuscript.

Fidelis Ewenitie Eki Udoko contributed to conception, design, analysis, interpretation, and critically revised the manuscript.

Nicholas F Schlecht contributed to acquisition, analysis, interpretation, and critically revised the manuscript.

Yana Bromberg contributed to conception, design, analysis, acquisition, interpretation, and critically revised the manuscript.

Nosayaba Osazuwa-Peters contributed to conception, design, analysis, acquisition, interpretation, and critically revised the manuscript.

Modupe O Coker contributed to conception, design, analysis, acquisition, interpretation, and critically revised the manuscript.

All authors gave their final approval and agree to be accountable for all aspects of the work.

