## Supplemental Materials for "Oral HPV and Dental Profiles in Mothers and Youth with or without HIV"

**Supplementary Materials**

***Random Forest and Boruta Models for oral HPV Prediction***

#### We built separate models for mothers and youth. Candidate predictors spanned demographics, sexual behavior, oral-health indices (DMFT, PUFA, OHIS, GIS), and immunologic markers (CD4, CD8, CD4:CD8). Categorical fields were encoded as factors; continuous fields were kept numeric. To minimize case loss in the youth model—where HPV positives were rare—missing numeric values were imputed with within-column medians and missing factor levels were set to “Unknown.” No scaling or resampling was applied.

#### **Random Forest (RF) classification**: RF models (R randomForest package; set.seed = 123) predicted HPV DNA detection (positive vs negative) using 1,000 trees (ntree = 1000) with default mtry (√p for classification). Model performance was summarized by out-of-bag (OOB) error rates. Variable importance was quantified with permutation-based mean decrease in accuracy (MDA), interpreted as the drop in OOB accuracy when a predictor is randomly permuted while all others are kept intact. Higher MDA indicates stronger association with the outcome. For presentation, importance scores were plotted and grouped by domain (Demographics, Sexual Behavior, Oral Health, Immunologic, Substance Use).

Given class imbalance (especially among youth), we report overall OOB error but emphasize variable-ranking stability rather than absolute accuracy. Partial-dependence plots were examined for the top predictors to visualize marginal associations.

#### **Feature-selection with Boruta**: To corroborate RF importance rankings, we applied the Boruta wrapper (R Boruta package; set.seed = 123). Boruta iteratively compares each real predictor’s RF importance to the maximum importance achieved by “shadow” (permuted) copies of the features, classifying variables as Confirmed, Tentative, or Rejected. We ran up to 100 iterations (maxRuns = 100) and then applied TentativeRoughFix to resolve tentative features.

#### **R environment:** Analyses were conducted in R using: randomForest, Boruta, ggplot2, dplyr, tidyr, forcats, cowplot, and patchwork. Exact package versions and seeds are listed in the analysis script.

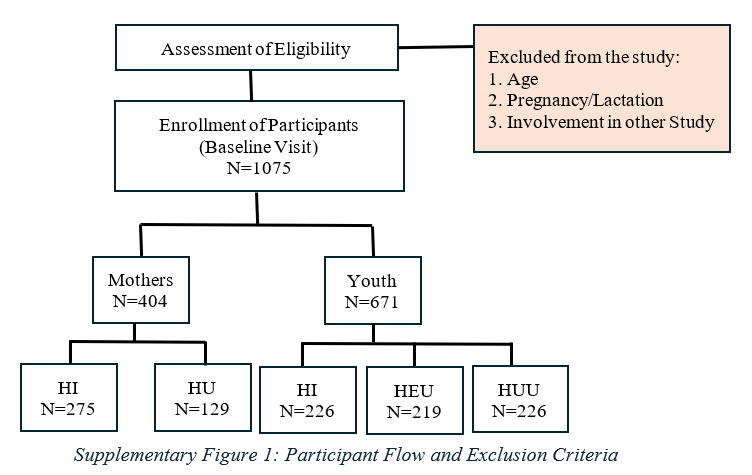

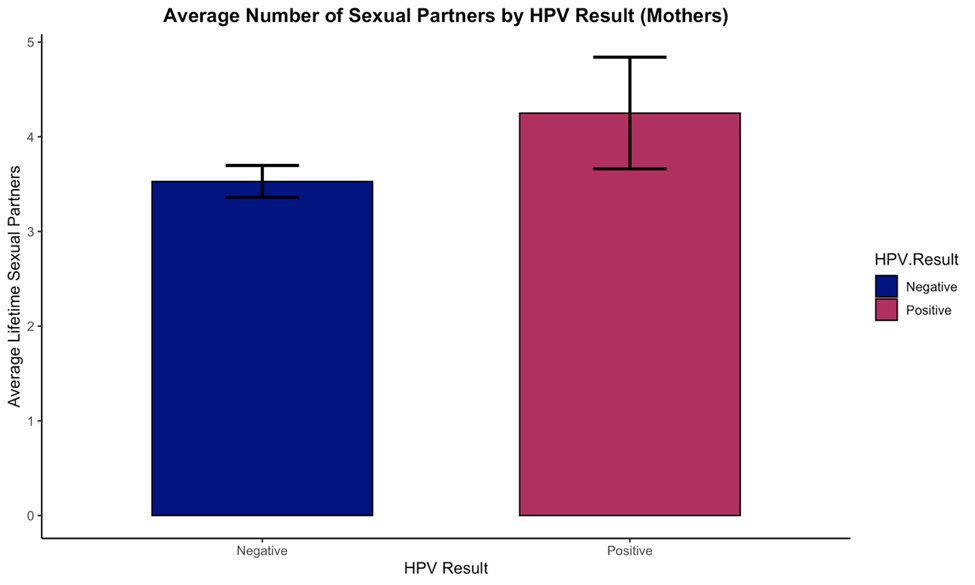

*Supplementary Figure 2: Average number of sexual partners for mothers stratified by HPV Status*

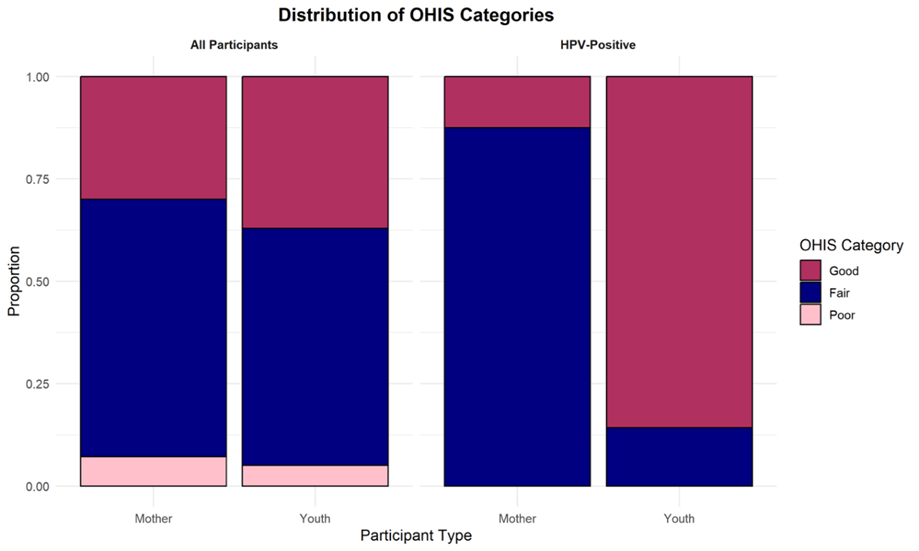

*Supplementary Figure 3: Oral Hygiene Index Simplified scores among mothers stratified by HPV Status*

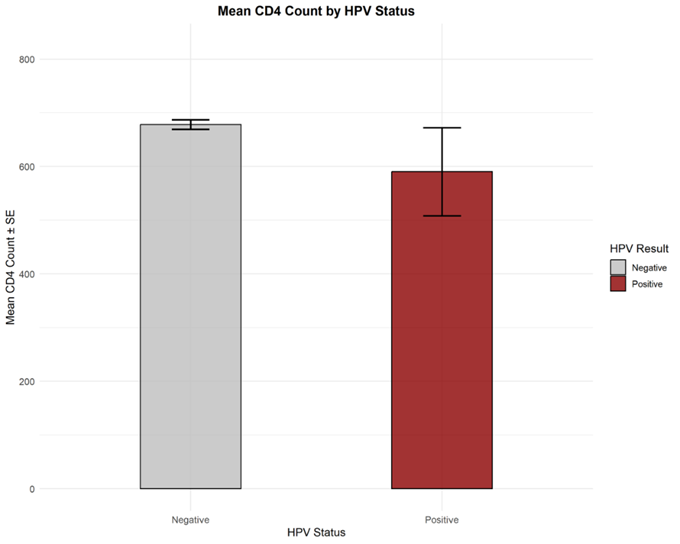

*Supplementary Figure 4: Average CD4 Count among Mothers based on HPV Status*

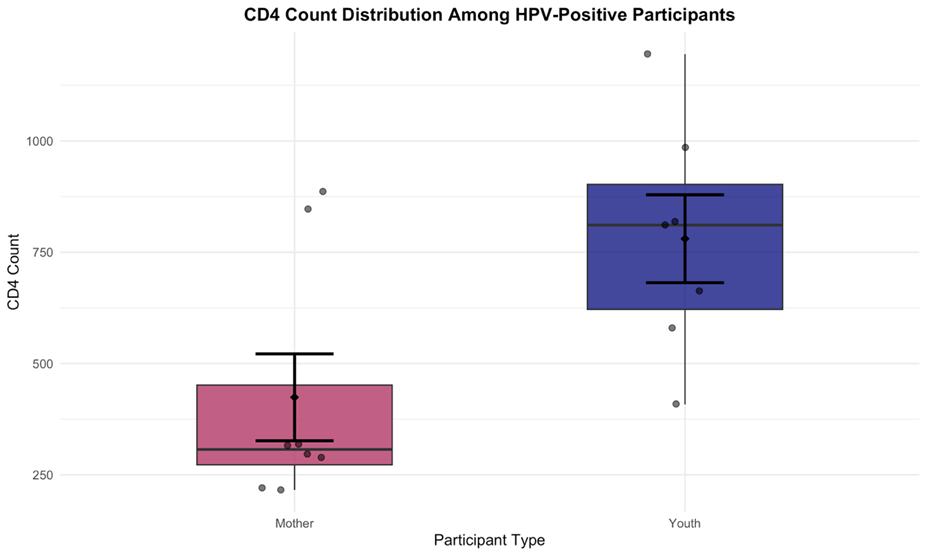

*Supplementary Figure 5: CD4 Count Distribution among Mothers and Youth with HPV DNA*

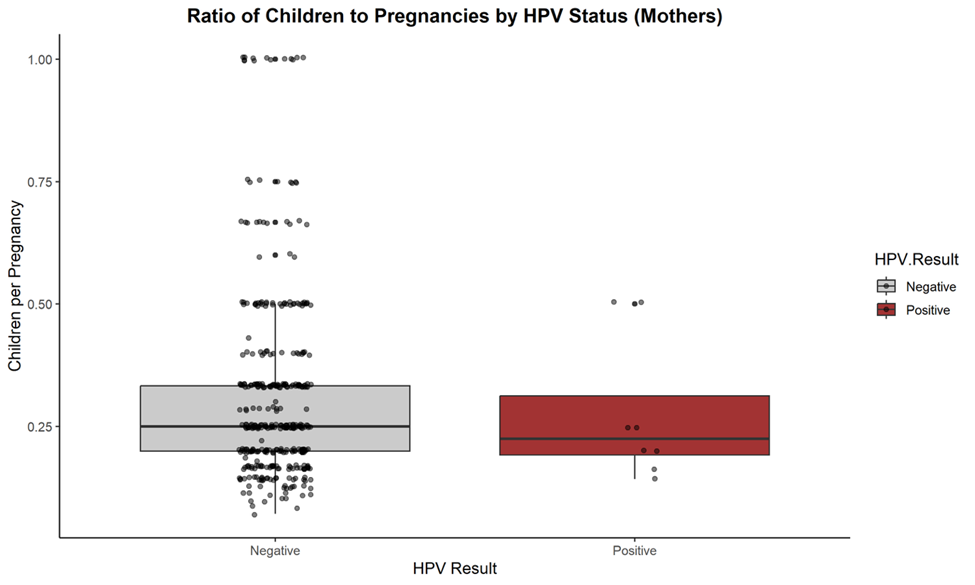

*Supplementary Figure 6: Parity among Mothers stratified by HPV Status*

| **Sample** | **Ct** | **HPV DNA** |
| --- | --- | --- |
| Y1 | 22.2 | 59 |
| Y2 | 22.5 | 39 |
| Y3 | 23.8 | 40 |
| Y4 | 25.9 | 16 |
| M1 | 29.3 | 56 |
| M2 | 30.9 | 35 |
| Y5 | 32.1 | 61 |
| M3 | 33.1 | 70 |
| M4 | 34.4 | 58 |
| M5 | 36.5 | 16 |
| Y6 | 37.5 | 66 |
| M6 | 38.3 | 35 |
| M7 | 38.5 | 16 |
| M8 | 38.8 | 11 |
| Y7 | 41.6 | 33 |

*Supplementary Table 1: Participants with oral rinse samples positive for HPV DNA (deidentified). Green: Low C(t) and reproducible; Yellow: High C(t) and reproducible.*
